# A comprehensive analysis of outcomes between COVID-19 patients with an elevated serum lipase compared to those with pancreatitis

**DOI:** 10.1101/2021.04.13.21252871

**Authors:** Petros C Benias, Sumant Inamdar, Diana Wee, Yan Liu, Jonathan M Buscaglia, Sanjaya K Satapathy, Arvind J Trindade, Northwell COVID-19 Research Consortium.

## Abstract

**Background and Aims:** COVID-19 patients may have asymptomatic hyperlipasemia without abdominal imaging findings or abdominal pain. In addition, primary and secondary pancreatitis have been described in COVID-19 patients. There is limited information on how the groups compare in outcomes. The aim is to compare outcomes among these groups.

**Methods:** This is a retrospective study from 12 hospitals within one healthcare system examining outcomes between hospitalized COVID-19 patients with a lipase <3x upper limit of normal (ULN), asymptomatic hyperlipasemia (>3x ULN), secondary pancreatitis (typical respiratory COVID-19 symptoms and found to have pancreatitis), and primary pancreatitis (presenting with pancreatitis).

**Results:** Of 11,883 patients admitted with COVID-19, 1,560 patients were included: 1,155 COVID-19 patients with a normal serum lipase (control group), 270 with an elevated lipase <3x ULN, 46 patients with asymptomatic hyperlipasemia with a lipase 3xULN, 57 patients with secondary pancreatitis, and 32 patients with primary pancreatitis. On adjusted multivariate analysis, the elevated lipase <3x ULN and asymptomatic hyperlipasemia groups had worse outcomes. The mortality was OR1.6 (95% CI 1.2-2.2) and 1.1 (95% CI 0.5-2.3), respectively. The need for mechanical ventilation was OR 2.8 (95% CI 1.2-2.1) and 2.8 (95% CI 1.5-5.2), respectively. Longer length of stay was OR 1.5 (95%CI 1.1-2.0) and 3.16 (95%CI 1.5-6.5), respectively.

**Conclusion:** COVID-19 patients with an elevated lipase< 3x ULN and asymptomatic hyperlipasemia have generally worse outcomes than those with pancreatitis. This could be attributed to extrapancreatic causes (liver failure, renal failure, enteritis, etc), which may signify a more severe course of clinical disease.

## INTRODUCTION

Gastrointestinal manifestations of the novel coronavirus disease (COVID-19) include nausea, vomiting, diarrhea, and increased liver function tests^1,2^. Recent studies suggest that COVID-19 can also present primarily as acute pancreatitis ^3–5^. The mechanism of injury in these cases is hypothesized to be via SARS-CoV-2 binding to islet cells of the pancreas that contain ACE2 receptors causing subsequent pancreatic injury ^5^.

While some patients present with clear symptoms of pancreatitis, others have been noted to simply have asymptomatic hyperlipasemia ^6^ with varying degrees of elevation of serum lipase. It is generally agreed upon that significant serum lipase elevation for a diagnosis of pancreatitis is three times the upper limit of normal^7^. Other organs can also secrete lipase such as the liver, kidney, and small intestine; although at generally lower serum concentrations than pancreatic lipase^8,9^. An elevated serum lipase may also reflect impaired clearance secondary to liver or renal failure^10^. Acute pancreatitis requires two of the following as per the revised Atlanta classification: 1) lipase greater than three times the upper limit of normal, 2) cross sectional imaging (computed tomography or magnetic resonance imaging) showing pancreatitis, and 3) characteristic upper abdominal pain at hospital admission ^7^.

It is unclear how outcomes (mortality, need for mechanical ventilation, and length of stay) compare in COVID-19 patients with elevated lipase <3x ULN, asymptomatic hyperlipasemia with a lipase >3xULN, and pancreatitis compared to COVID-19 patients with a normal lipase. The aim of our study is to compare these outcomes among these groups.

## METHODS

This is a retrospective observational cohort study of COVID-19 patients 18 years or older admitted to twelve hospitals within the Northwell Health System from March 1, 2020-June 1, 2020 during the COVID-19 pandemic in New York. Institutional Review Board approval (IRB #20-0200, Registry of patients who are presenting under the suspicion of COVID-19) was obtained for this study by the Northwell Health Feinstein Institute for Medical Research, Office of the Human Research Protection Program (Hallie Kassan, MD; Director). Patients charts were searched for based on lipase values, cross sectional imaging (computed tomography or magnetic resonance imaging) showing pancreatitis, or charts being coded for acute pancreatitis.

The following patient groups were compared: COVID-19 patients with a normal lipase, COVID-19 patients with an elevated lipase but less than 3x ULN, COVID-19 patients with a lipase >3xULN but without pancreatitis (no imaging concerning for pancreatitis or abdominal pain), COVID-19 patients with primary pancreatitis (presenting with acute pancreatitis and subsequently found to have COVID-19), and COVID-19 patients with secondary pancreatitis (patients presenting with respiratory COVID-19 symptoms (e.g. shortness of breath and fever) and found to have pancreatitis). The primary pancreatitis group has been previously reported^11^. Patient charts with a lipase >3xULN were subsequently individually reviewed to determine the etiology of pancreatitis if applicable, why a serum lipase was ordered, and to confirm group categorization. Patients with acute pancreatitis met the revised Atlanta classification (as previously defined). The primary outcomes of mortality, lengths of stay, and need for mechanical ventilation were compared between the groups.

Univariate and bivariate analysis was performed using students’ t-test or ANOVA for comparison of continuous variables, and Chi square test for comparison of categorical variables. Multivariate analysis was performed using proc logistic and the model controlled for diabetes mellitus, gender, hypertension, congestive heart failure, chronic obstructive pulmonary disease, and Charlson comorbidity index (CCI). SAS, Version 9.4 (SAS Institute, Cary, NC) was used to perform all analysis. The cohort admitted with primary pancreatitis has been previously compared to patients admitted with pancreatitis without COVID-19 ^3^.

## RESULTS

Of 11,883 patients admitted with COVID-19, 1,560 patients were included: 1,155 COVID-19 patients with a normal serum lipase (control group), 270 with an elevated lipase <3x ULN, 46 patients with asymptomatic hyperlipasemia >3xULN, 57 patients with secondary pancreatitis, and 32 patients with primary pancreatitis. On manual chart review, all 46 patients with asymptomatic hyperlipasemia had a serum lipase ordered, as it was part of a critical care serum panel for critically ill patients. There was no documentation of abdominal pain or suspicion for pancreatitis. In patients with primary and secondary pancreatitis, a serum lipase was ordered for the clinical suspicion of pancreatitis.

Patient characteristic can be found in Table 1. The primary pancreatitis group was statistically younger compared to controls (53 vs 63; p=0.004), but the rest of the groups were similar in age. There were a higher percentage of females in the lipase <3xULN and the secondary pancreatitis group. There were racial differences between the lipase <3xULN group and controls, but not among other groups. The lipase <3xULN group had a higher proportion of patients with diabetes compared to the control group. The lipase <3xULN and primary pancreatitis groups had a higher proportion of ptients with hypertension compared to the controls. The primary pancreatitis group had lower Charlson Comorbidity Index (1-2), compared to the other groups..

**Table 1:**
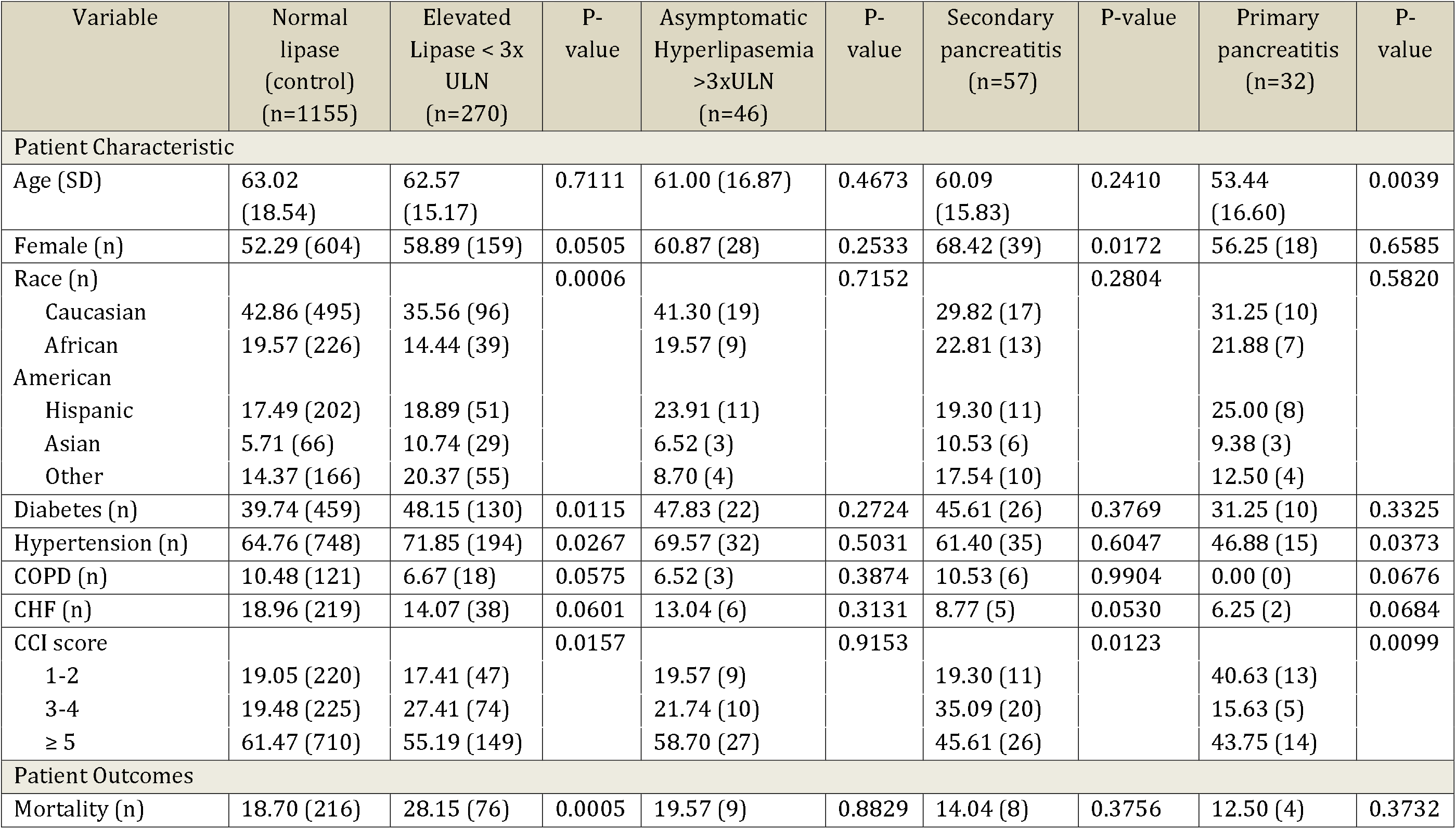
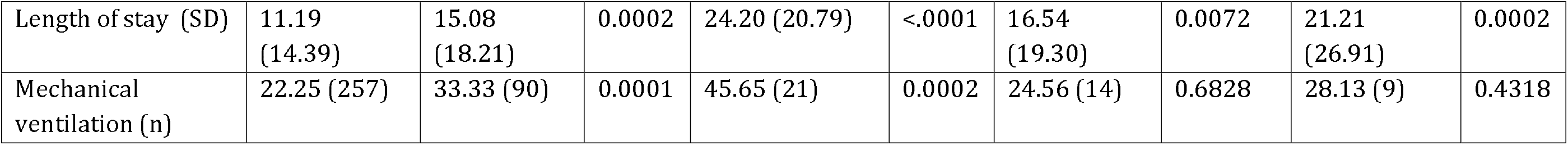
Patient Characteristics

The unadjusted and adjusted univariate and multivariate analysis can be found in Table 2. On adjusted multivariate analysis, the elevated lipase <3x ULN and asymptomatic hyperlipasemia with a lipase > 3xULN groups had generally worse outcomes compared to the secondary and primary pancreatitis groups. Odds ratios (OR) for mortality were as follows: OR 1.6 (95% CI 1.2-2.2) for the elevated lipase <3x ULN group, OR 1.1 (95% CI 0.5-2.3) for the asymptomatic hyperlipasemia group, versus OR 0.70 (95% CI 0.32-1.5) and OR 0.73 (95% CI 0.25-2.17) for the secondary pancreatitis and the primary pancreatitis groups. Need for mechanical ventilation was also greater in these two groups: OR 2.8 (95% CI 1.2-2.1) for the elevated lipase <3X ULN group and OR 2.8 (95% CI 1.5-5.2) for the asymptomatic hyperlipasemia group, versus OR 0.95 (95% CI 0.5-1.8) and OR 1.3 (95% CI 0.6-3.0) for the secondary and primary pancreatitis groups, respectively. Similarly, longer length of stay was more likely in the two groups without Atlanta criteria defined pancreatitis: OR 1.5 (95% CI 1.1-2.0) and OR 3.2 (95% CI 1.5-6.5) for the elevated lipase <3x ULN group and asymptomatic hyperlipasemia group, respectively; versus OR 1.3 (95% CI 0.77-2.3) and OR 2.2 (95% CI 1.0-4.8) for secondary and primary pancreatitis groups, respectively.

**Table 2:**
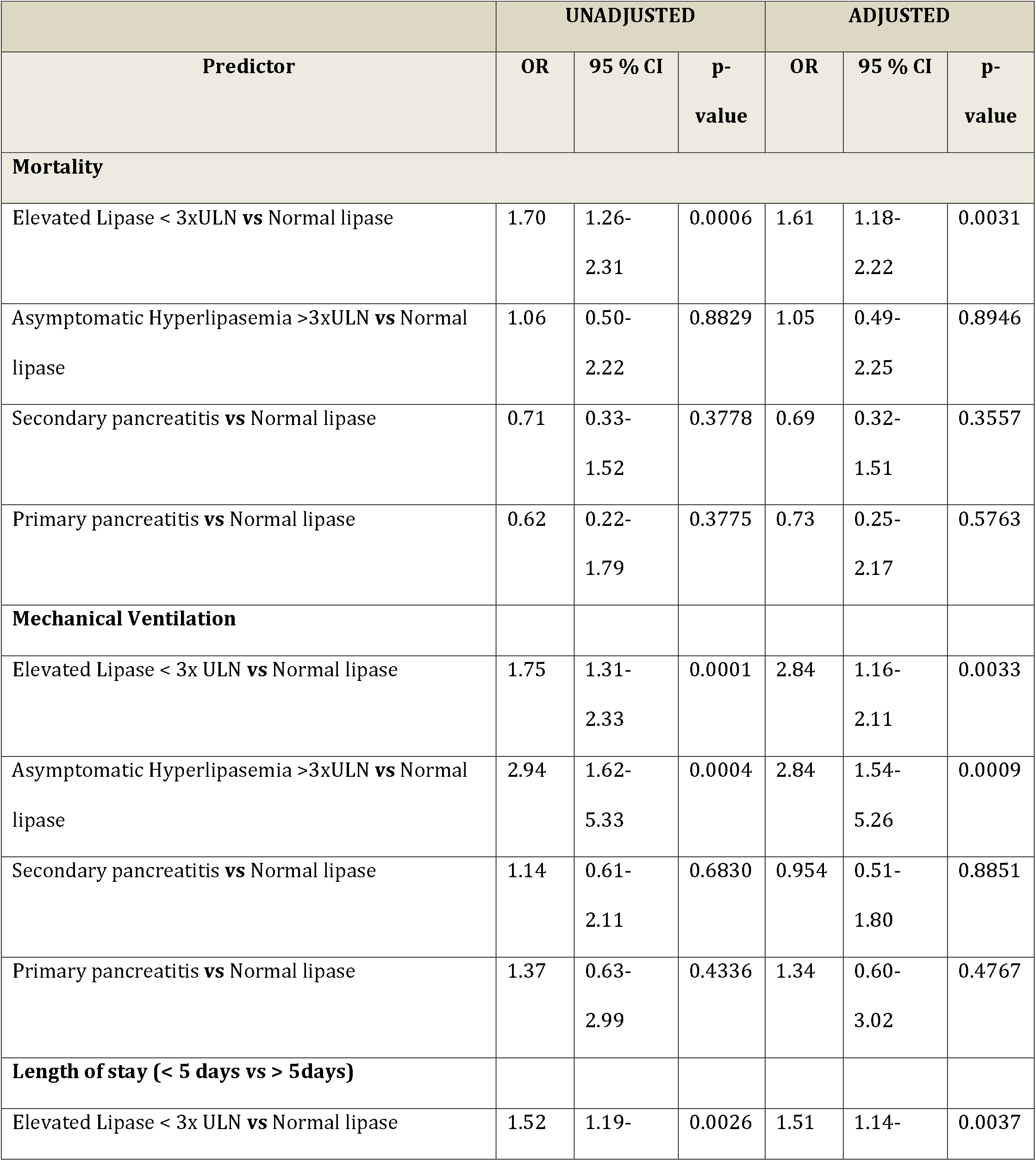
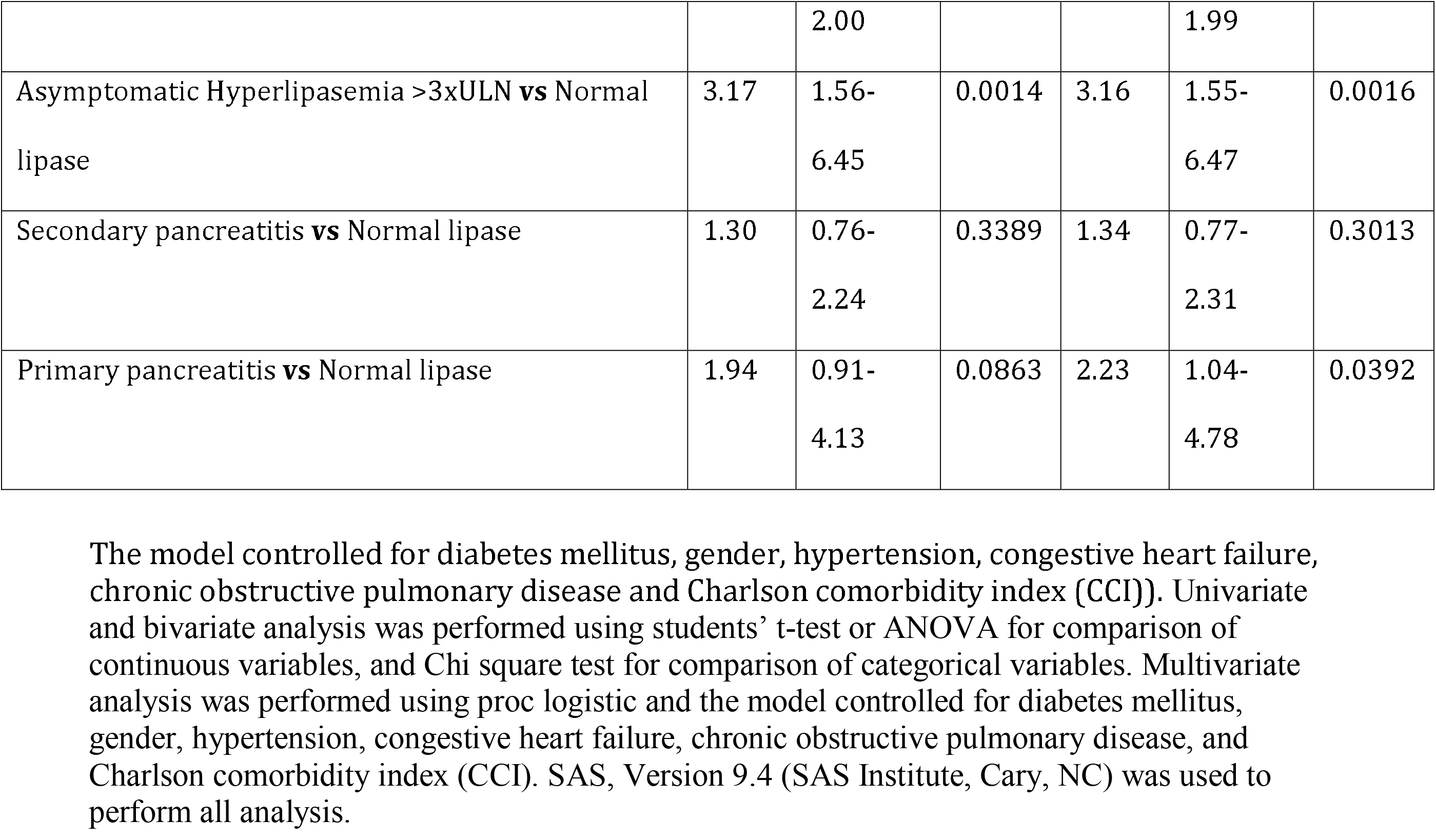
Unadjusted and Adjusted Odds Ratios for Outcomes of the Study

## DISCUSSION

This study examined the difference in outcomes between hospitalized COVID-19 patients with an elevated lipase <3x ULN, asymptomatic hyperlipasemia with a lipase > 3xULN, secondary pancreatitis, and primary pancreatitis. These groups were compared to COVID-19 patients with a normal lipase. After controlling for many variables, the groups with an elevated lipase <3x ULN and asymptomatic hyperlipasemia with a lipase > 3xULN fared worse than those with pancreatitis. The ORs of mortality for the elevated lipase <3x ULN and asymptomatic hyperlipasemia with a lipase >3xULN were above 1, although the asymptomatic hyperlipasemia p-value was not significant. This is likely due to the small nature of the group (n=46) compared to the elevated lipase <3xULN group (n=270). The pancreatitis groups ORs were well under 1 for mortality. The need for mechanical ventilation was also worse for the elevated lipase <3x ULN and asymptomatic hyperlipasemia with a lipase >3xULN groups (Both with ORs 2.84 and statistically significant). Finally, both groups had longer length of stays, although the primary pancreatitis group also had a longer LOS.

Pancreatic lipase is stored in the pancreas at concentrations 20,000 that of serum concentrations^8^. During pancreatitis, lipase is released in large amounts into the circulation^12^. Serum lipase can be elevated in a number of other conditions such as cirrhosis, renal failure, cancer, enteritis, colitis, infections, trauma, hyperglycemia^8^. However these elevations are not as high compared to pancreatitis due to storage capacity of lipase in the pancreas^8,9^. Thus the COVID-19 group with an elevated lipase <3xULN and the COVID-19 group with asymptomatic hyperlipasemia (no imaging concerning for pancreatitis or abdominal pain) likely have an elevated lipase due to a nonpancreatic source. Thus, the hyperlipasemia and worse outcomes in these groups could be attributed to nonpancreatic injury or impaired clearance of pancreas lipase by the liver, kidney, etc. Given the many causes of nonpancreatic hyperlipasemia, the only true way of knowing the etiology would be to fractionate the lipase, which is beyond the scope of this retrospective study. In addition it would be hard to control for all the sources of nonpancreatic hyperlipasemia in this study.

Our study has notable strengths. Our study is the first to compare COVID-19 patients with pancreatitis to COVID-19 patients with asymptomatic hyperlipasemia to 3xULN and those with an elevated lipase <3x ULN using a control of COVID-19 patients with a normal lipase. There have been numerous reports on hyperlipasemia, and thus we feel this comprehensive analysis including the whole spectrum of possible elevations of serum lipase provides a clearer picture on the topic. In addition, we do compare patients with pancreatitis presenting as COVID-19 versus COVID-19 patients presenting with respiratory symptoms found to have pancreatitis. Although this is a subtle clinical difference, it was unknown if the outcomes are different in this patient population. Finally, the study is from a diverse and large health system in New York from Long Island, Manhattan, Queens, and Staten Island. This increases the generalizability of the study.

Our study has limitations. First, it is retrospective in design and thus has its inherit limitations. Second, a serum lipase was ordered as part of a critical care serum panel for ill patients and thus may bias towards sicker patients. Finally the hypothesis that the COVID-19 group with lipase <3xULN and the COVID-19 group with asymptomatic hyperlipasemia (no imaging concerning for pancreatitis or abdominal pain) likely have an elevated lipase due to a nonpancreatic source can only truly be proven by fractionating the lipase which was not done in these patients as it is not part of standard clinical care.

In conclusion, this study attempts to provide clarity regarding hyperlipasemia in patients with COVID-19. We found that COVID-19 patients with a lipase <3x ULN and asymptomatic hyperlipasemia with a lipase > 3xULN generally have worse outcomes compared to those with pancreatitis. Future studies from similar large health systems may be need to confirm our findings.

## Data Availability

Data is available upon request

## Acknowledgements

None

## Notes

### Competing Interest Statement

PCB: Consultant for Olympus America, Apollo Medical, FujiFilm, and Boston Scientific
JB: Consultant and speaker for Abbvie Inc.
AJT: Consultant for Olympus America and Pentax Medical
Research Support form Ninepoint Medical

### Funding Statement

This project was unfunded.

### Author Declarations

Institutional Review Board approval (IRB #20-0200, Registry of patients who are presenting under the suspicion of COVID-19) was obtained for this study by the Northwell Health Feinstein Institute for Medical Research, Office of the Human Research Protection Program (Hallie Kassan, MD; Director)

